# Automatic Breast Cancer Cohort Detection from Social Media for Studying Factors Affecting Patient Centered Outcomes

**DOI:** 10.1101/2020.05.17.20104778

**Authors:** Mohammed Ali Al-Garadi, Yuan-Chi Yang, Sahithi Lakamana, Jie Lin, Sabrina Li, Angel Xie, Whitney Hogg-Bremer, Mylin Torres, Imon Banerjee, Abeed Sarker

## Abstract

Breast cancer patients often discontinue their long-term treatments, such as hormone therapy, increasing the risk of cancer recurrence. These discontinuations may be caused by adverse patient-centered outcomes (PCOs) due to hormonal drug side effects or other factors. PCOs are not detectable through laboratory tests, and are sparsely documented in electronic health records. Thus, there is a need to explore complementary sources of information for PCOs associated with breast cancer treatments. Social media is a promising resource, but extracting true PCOs from it first requires the accurate detection of breast cancer patients. We describe a natural language processing (NLP) architecture for automatically detecting breast cancer patients from Twitter based on their self-reports. The architecture employs breast cancer related keywords to collect streaming data from Twitter, applies NLP patterns to pre-filter noisy posts, and then employs a machine learning classifier trained using manually-annotated data (n=5019) for distinguishing firsthand self-reports of breast cancer from other tweets. A classifier based on bidirectional encoder representations from transformers (BERT) showed human-like performance and achieved F_1_-score of 0.857 (inter-annotator agreement: 0.845; Cohen’s kappa) for the positive class, considerably outperforming the next best classifier—a deep neural network (F_1_-score: 0.665). Qualitative analyses of posts from automatically-detected users revealed discussions about side effects, non-adherence and mental health conditions, illustrating the feasibility of our social media-based approach for studying breast cancer related PCOs from a large population.

## 1 Introduction

### 1.1 Background

Women with breast cancer comprise the largest group of cancer survivors^¶^ in high-income countries such as the United States, particularly due to the availability of advanced treatments (*e.g*., hormone therapy) that have significantly reduced mortality rates. Due to the treatment-driven increased life expectancy of breast cancer survivors, their physical and psychological well-being are regarded as important patient-centered outcomes (PCOs), specifically among younger patients. Breast cancer patients often suffer from various treatment-related side effects and other negative outcomes, which range from short-term pain, nausea and fatigue, to lingering psychological dysfunctions such as depression, anxiety, and suicidal tendency. Consequently, one-third to half of young breast cancer patients discontinue their treatments, such as endocrine therapy, thus increasing the risk of cancer recurrence and therefore of death [7, 8]. In addition, nonadherence to prescribed therapy is associated with poor quality of life, more physician visits and hospitalizations, and longer hospital stays [9].

PCOs, including treatment-related side-effects, are not captured in laboratory or diagnostic tests, but are gathered through patient communications. Sometimes these outcomes are captured as free text in clinical narratives written by caregivers. PCOs documented in this manner, however, are often subject to biases and incompleteness of data in the Electronic Health Records (EHR). In many cases PCOs are not documented at all. We demonstrated the under-documentation of PCOs of oncology patients in EHRs in a recent study [2]. Specifically, with the approval of Stanford Institutional Review Board (IRB), we deployed a simple rule-based NLP pipeline for breast cancer, which searched for documentation of physical and mental PCOs affecting patient well-being in EHRs. Physical PCOs (type 1 PCOs) consisted of pain, nausea, hot flush, fatigue, while mental PCOs (type 2) included anxiety, depression and suicidal tendency. On 100 randomly selected clinical notes of breast cancer patients, the model achieved 0.9 F_1_-score when validated against manually-labeled ground truth. We applied the validated model on the Stanford breast cancer dataset (Oncoshare), which contains an assortment of clinical notes (*e.g*., progress notes, oncology notes, discharge summaries, nursing notes) associated with 8,956 women diagnosed with breast cancer from 2008 to 2018. As depicted in Table 1, only 8% of clinical notes and 12% of progress notes contained any documentation (affirm/negation) of PCOs. Importantly, for as many as 30% of breast cancer patients, there were no documented PCOs at any time point at all.

**Table 1.**
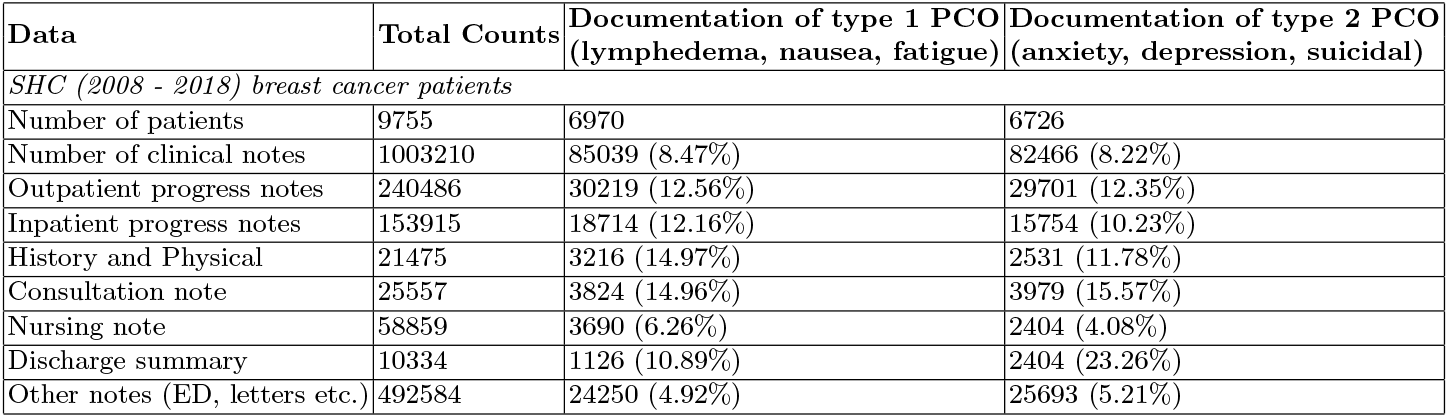
Results of patient-centered outcome extraction from clinic notes of Stanford Breast Cancer Cohort (2008 - 2018).

The under-documentation of PCOs acts as a limiting factor to study the long-term treatment outcomes of young breast cancer patients. Most of the past studies focusing on PCOs have either relied on only small populations of clinical trial patients or analyzed short-term side effects collected during frequent clinic visit periods. Another important limiting factor to understanding the outcomes that matter to patients is that studies focusing on EHRs only capture clinical information, not other relevant factors and patient characteristics that influence their long- and short-term outcomes. Some studies have investigated the feasibility of monitoring patient-reported outcomes (PROs) among oncology patients using sources other than EHRs, such as web portals, mobile applications and automated telephone calls, and their findings suggest that monitoring PROs outside of clinic visits may be more effective and reduce adverse outcomes. However, engaging oncology patients in such routine monitoring activities is extremely resource intensive (expensive) and they only enable the collection of limited information from homogeneous cohorts. Given the under-documentation in EHRs and the laborious process of conducting patient surveys, there is a need to identify complementary sources of information for PCOs associated with breast cancer patients/survivors, and to develop new strategies for capturing diverse patient-level and population-level health-related outcomes.

One promising, albeit challenging, source of information for population-level breast cancer PCOs/PROs is social media. Several studies, including our own, have utilized social media to identify large cohorts of users with common health-related conditions, and then mine relevant longitudinal information about the cohorts using NLP methods. For example, in our past research, we showed that carefully-designed NLP pipelines can be used to discover cohorts of pregnant women [11] or patients suffering from opioid use disorder [6] from social media, and then mine important information from their social media posts (*e.g*., medication usage and recovery strategies). For cancer, studies have investigated the role of social media platforms for tasks such as spreading breast cancer awareness, health promotion, and cancer prevention [1, 3]. However, to the best of our knowledge, no past research has attempted to accurately detect cancer cohorts from social media to study long-term cohort-specific information at scale.

### 1.2 Objectives

We had the following 3 specific objectives for this study, each dependent on the previous one:

a. Assess if breast cancer patients discuss personal health-related information on Twitter, including the self-reporting of their positive breast cancer diagnosis/status.
b. Develop a social media mining pipeline for detecting self-reports of breast cancer using NLP and machine learning methods from Twitter (the primary aim of the paper).
c. Gather longitudinal information from the profiles of the automatically-detected users, and qualitatively analyze the information to ascertain if long-term research can be conducted on this cohort.

## 2 Materials and Methods

### 2.1 Data and Annotation

We collected data from Twitter using keywords and hashtags via the public streaming application programming interface (API). We used four keywords: (i) cancer, (ii) breast cancer, (iii) tamoxifen, (iv) survivor, and their hashtag equivalents. An inspection of Twitter data retrieved by these keywords showed that while there are many health-related posts from real breast cancer patients, they were hidden within large amounts of noise. Table 2 shows examples of tweets mentioning these keywords, including breast cancer self-reports (category: **S**), and tweets that were not relevant (category: **NR**). We filtered out most of the irrelevant tweets by employing several simple rule- and pattern-matching methods, only keeping tweets that matched the patterns, which were as follows:

**Table 2.** Sample tweets from keyword-based retrieval of data from Twitter. Tweets have been modified to preserve anonymity. ‘*’ - tweet filtered by pattern-matching; ‘**’ - tweet not filtered by pattern-matching (requiring supervised classification).

| Tweet | Pattern/Keyword Match | Category |
| --- | --- | --- |
| I am blessed. I know this. As one of the lucky ones, my breast cancer was caught early on. Almost five years ago. @USERNAME URL #survivor #amwriting #writingcommunity #writerlift screenwriters | breast & cancer & survivor | S |
| It’s damn hard to fight cancer when you cold, hungry & live with constant financial stress. | cancer* | NR |
| Check out Shelby J’s latest single regarding her recent struggle with breast cancer and what sustained her throughout. #Survivor #EarlyDetectionSavesLives #MusicMonday | breast & cancer & survivor** | NR |
| Im officially a 16 year breast cancer survivor , mammogram came back all clear no evidence of recurring disease. So grateful | breast & cancer & survivor | S |

**Table 3.**
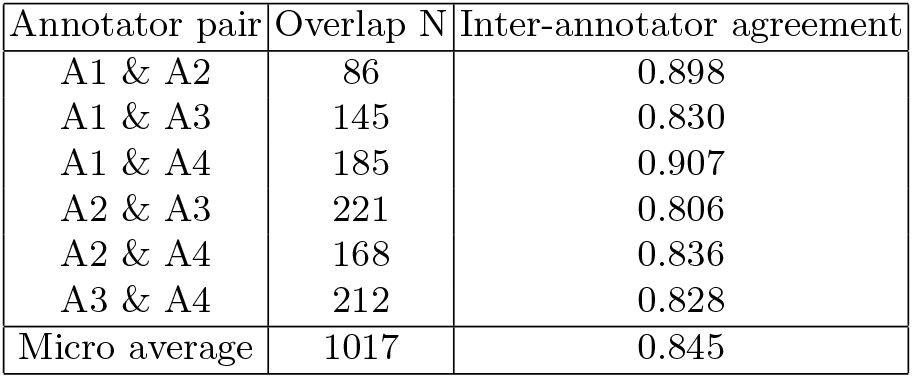
Pair-wise IAAs, numbers of overlapping tweets, and overall micro average.

| Annotator pair | Overlap N | Inter-annotator agreement |
| --- | --- | --- |
| A1 & A2 | 86 | 0.898 |
| A1 & A3 | 145 | 0.830 |
| A1 & A4 | 185 | 0.907 |
| A2 & A3 | 221 | 0.806 |
| A2 & A4 | 168 | 0.836 |
| A3 & A4 | 212 | 0.828 |
| Micro average | 1017 | 0.845 |

– Tweet contains [#]breast & [#]cancer & [#]survivor; OR
– Tweet contains [#]breastcancer & #survivor; OR
– Tweet contains [#]tamoxifen AND ([#]cancer OR [#]survivor)
– Tweet contains a personal pronoun (*e.g*., ‘my’, ‘I’, ‘me’, ‘us’) AND [#]breast & [#]cancer

These patterns were developed via a brief manual analysis of Twitter chatter using the website (i.e., the search option). From Table 2, we see that the pattern-based filter does not remove all irrelevant tweets. To fully automate the detection and collection of a Twitter breast cancer cohort, it is necessary to detect self-reports with higher accuracy. Therefore, we employed supervised classification, similar to our past research focusing on Twitter and a pregnancy cohort [11]. We chose a random sample of the pre-filtered tweets for manual annotations. We excluded duplicate tweets, retweets and tweets shorter than 50 characters. Four annotators performed the annotation of tweets, with a random number of overlapping tweets between each pair of annotators. Each tweet was labeled as one of three classes-(i) self-report of breast cancer (S), (ii) report of breast cancer of a family member or friend (F), or (iii) not relevant (NR). We computed pair-wise inter-annotator agreements using Cohen’s kappa [4]. Since we were only interested in first person self-reports of breast cancer for this study, we combined classes F and NR for the supervised machine learning experiments.^‖^

### 2.2 Supervised Classification

We experimented with multiple supervised classification approaches and compared their performances on the same dataset. These approaches were naive Bayes (NB), random forest (RF), support vector machine (SVM), deep neural network (NN), and a classifier based on bidirectional encoder representations from transformers (BERT). For the NB, RF, and SVM classifiers, we pre-processed by lowercasing, stemming, removing URLs, usernames, and non-English characters. Following the pre-processing, we converted the text into features: n-grams (contiguous sequences of n words ranging from 1 to 3), and word clusters (a generalized representations of words learned from medication-related chatter collected from Twitter) [12]. For these classifiers, we used *count vector* representations—each tweet is represented as a sparse vector whose length is the size of the entire feature-set/vocabulary and each vector position represents the number of times a specific feature (*e.g*., a word or bi-gram) appears in the tweet. In addition to being sparse (*i.e*., most of the vector numbers are 0), these count-based representations do not capture word meanings or their similarities. For instance, the terms ‘bad’ and ‘worst’ will be represented by orthogonal vectors. Word embedding based representations such as GloVe [10] capture word meanings and we used them for the NN classifier. However, such representations do not capture contextual differences in the meanings of words.

Transformer-based approaches, such as BERT, encode contextual semantics at the sentence or word-sequence level, and have vastly improved the state-of-the-art in many NLP tasks [5]. BERT-based classifiers had not been previously used for health cohort detection from Twitter, and in this study, we used the *BERT large* model [5] which consists of 16 layers (transformer blocks), 1024 hidden size 16 attention heads with total of 340M parameters. The tweets are converted into the BERT model, which captures contextual meanings of character sequences. Following vectorization, a neural network (dense layer) with a softmax activation is used to predict whether the tweets is (NR or S).

### 2.3 Post-classification Analyses

Following the classification experiments, we conducted manual analyses to (i) study causes of classification errors, (ii) analyze the association between training set size and classification performance for all classifiers, and (iii) verify if the users detected by the classification approach discussed factors that influenced PCOs on Twitter. For (i) we manually reviewed a sample of the misclassified tweets to identify potential patterns. For (ii), our objective was to assess if the number of tweets required to obtain acceptable classification performance was practical and feasible. We drew stratified samples of the training set consisting of 20%, 40%, 60% and 80% of the set, and computed the F_1_-scores over the same test set. For (iii), we collected, via the API, the past posts of a subset of automatically-detected breast cancer positive users, and then qualitatively analyzed them. We used simple string-matching to identify potentially relevant tweets.

## 3 Results

### 3.1 Annotation and Supervised Classification Results

We annotated a total of 5,019 unique tweets (training: 3513; validation: 302; evaluation: 1204). 3736 (74%) tweets belonged to the NR class (training: 2615; validation: 225; test: 896) and 1283 (26%) belonged to the S class (training:898; validation: 77; test: 308). Micro-average of the pair-wise agreements among all annotators was 0.845 (Cohen’s κ) [4], which represents significant agreement [13]. Table 3.1 presents IAA for each pair of annotators.

Table 4 shows the performances of the learning algorithms on the held-out test set. The BERT-based classifier yields the highest F _1_-score for class S (0.857), significantly outperforming the other classifiers.

**Table 4.**
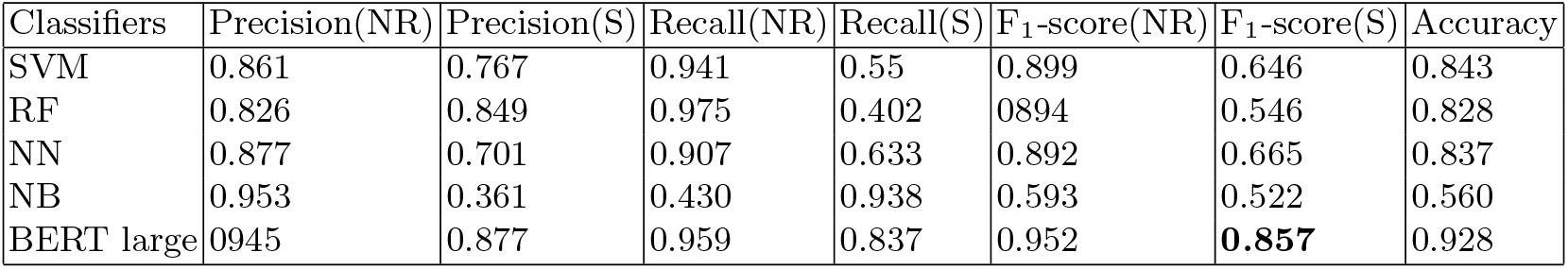
Performances of learning models in terms of class-specific recall, precision, F_1_-scores, and overall accuracy. Best F_1_-score on the 1 class is shown in bold. tables.

| Classifiers | Precision(NR) | Precision(S) | Recall(NR) | Recall(S) | F<sub>1</sub>-score(NR) | F<sub>1</sub>-score(S) | Accuracy |
| --- | --- | --- | --- | --- | --- | --- | --- |
| SVM | 0.861 | 0.767 | 0.941 | 0.55 | 0.899 | 0.646 | 0.843 |
| RF | 0.826 | 0.849 | 0.975 | 0.402 | 0.894 | 0.546 | 0.828 |
| NN | 0.877 | 0.701 | 0.907 | 0.633 | 0.892 | 0.665 | 0.837 |
| NB | 0.953 | 0.361 | 0.430 | 0.938 | 0.593 | 0.522 | 0.560 |
| BERT large | 0.945 | 0.877 | 0.959 | 0.837 | 0.952 | **0.857** | 0.928 |

### 3.2 Post Classification Analyses Results

Classification error analyses:

As per our analysis, the possible reasons for misclassification could be attributed to factors that are common with social media data, primarily the lack of context, ambiguous references, and the use of colloquial language. The following following tweets are classified by the annotator as S, but BERT misclassified them:

> Tweet-1: *“we are sisters in this breast cancer club we never wanted to join. bless you my friend. you are an inspiration to all of us.”*
>
> Tweet-2: *“when the breast cancer center calls and asks you to donate for the patients’ medication and you’re just like ”i can barely afford my own”*

#### Learning curve at different training data sizes

Figure 1 shows the classifier performances at different training data sizes with increments of 20% of the full training set. From the figure, we see that the BERT-based classifier shows remarkable performance even at small training set sizes. However, the performance of this classifier does not improve further as more training data is added.

**Fig. 1.**
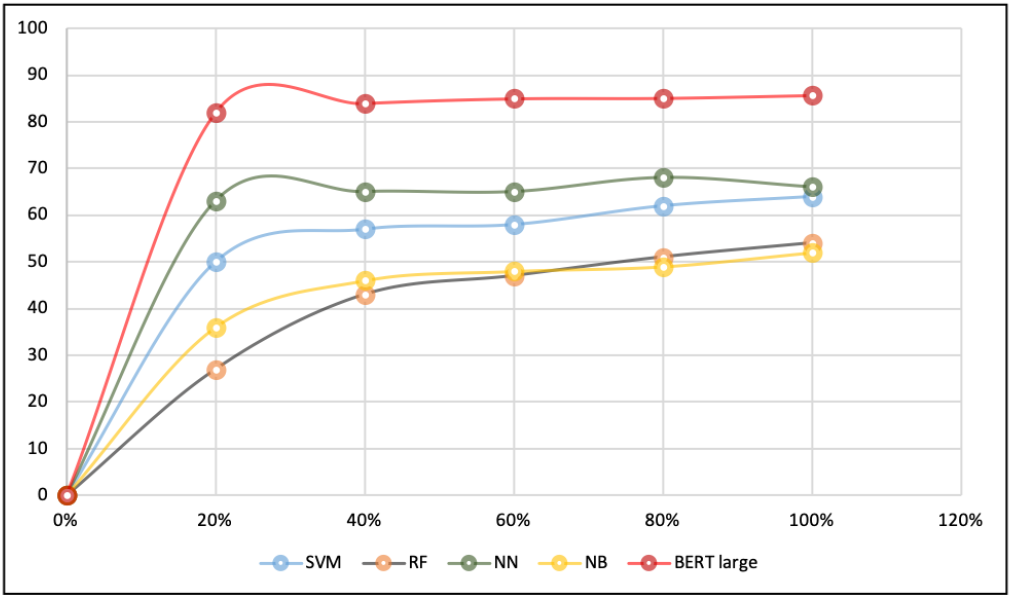
Classifier performances at different different training set sizes.

#### Content exploration

We found many informative tweets that covered a wide variety of health-related, and potentially cancer-related, information. Table 5 presents some examples of tweets that were potentially relevant to the users’ PCOs. A number of users reported that they suffered from anxiety/depression, although it was not immediately clear how their mental health conditions were related their cancer diagnoses and treatments. Similarly, users report experiencing or worrying about the side effects of prescribed medications, including Tamoxifen, and their intentions to not adhere to the treatment. These tweets could provide crucial information about how these survivors cope with their treatment and medications, complementing their EHRs.

**Table 5.** Sample posts that are relevant to the users’ health conditions, collected from the timelines of automatically-detected users. The posts were manually curated and categorized. URLs and emoji’s have been removed; usernames have been anonymized.

| # | Tweet | Comment |
| --- | --- | --- |
| # 1 | Sooooo..... the doc put me on an anxiety/anti-depression med the other day (cuz cancer is still a b*tch). She told me to take in the morning. Uh no. I’ve been asleep for 2 days almost. Taking that joint at night. | Mental health issues |
| # 2 | my #mentalhealth suffered unnecessarily and drastically due to #thyroid medications that didn’t work for me, for my body. even when #hypothyroid (on paper) is treated it can make you feel even more unwell. keep asking for help from new medical professionals until one listens. | Mental health issues |
| # 3 | Here we are at my Oncology follow up appointment. I didnt really get on with the tablets prescribed for hot flushes. They made me so sleepy I felt like a zombie and a lower mood than usual so I stopped them. Hopefully get echocardiogram results today too | Side effects, nonadherence intention |
| # 4 | I’m learning something new every day about my #breastcancer. While seeing the oncologist yesterday, I said I know if I stay on my 5 year hormone therapy plan, there is a 9% chance of recurrence. So I asked what if I stop taking the medicine so I no longer have joint pain... | Side effects |
| # 5 | New drug today Docetaxel. Not got my usual anti sickness prescribed so I’m feeling quite nervous about how it’s going to take me I was vomiting on the EC treatment. But on the positive this is number 5 of 8. #breastcancer #chemotherapy | Side effects |
| # 6 | And Im having a mentally poor day. For all its benefits in preventing #breastcancer recurrence, I think I am going to have to stop taking #Tamoxifen I have a review at the hospital shortly to discuss. Yes, I am grateful that this drug is available but the quality of life is poor | Side effects, nonadherence intention |
| # 7 | Another night, another with lack of sleep. How Im supposed to continue getting by on 3-4hrs sleep every night is beyond me and definitely contributing to my emotional state of mind. I havent had one night since pre #breastcancer where Ive slept all night #mentalhealth #tamoxifen | Mental health issues, side effects |
| # 8 | The prize for finishing chemo is taking a drug that can cause uterine cancer. #oneroundleft #breastcancer #tamoxifen | Side effects |

## 4 Discussion

The capability to detect self-reports of breast cancer very accurately is a necessary condition for utilizing Twitter to study PCOs associated with treatment, and our approach has produced promising results. The transformer-based classifier (BERT), is capable of producing performances that far outperform traditional approaches. Thus, our study demonstrates that it is indeed possible to build a large breast cancer cohort from Twitter via an automatic NLP pipeline.

Manual annotation of data is a very time-consuming task and the need to annotate large numbers of samples for supervised classification often act as a barrier to practical deployment. Our experiments show that the BERT-based model overcomes this obstacle, making full automation feasible. However, we also discovered that it is difficult to raise the performance of this classifier simply by annotating more data. Despite the context-incorporating sentence vectors that are used for BERT, the model still lacks the ability to infer meanings that are typically evident to humans. Also, our annotators benefited from implicit knowledge of the topic and additional contextual cues, which the transformer-based model is not able to capture. In the future, it will be important to study how such implicit information may be encoded in numeric vectors.

## 5 Conclusion

We investigated the potential of using Twitter as a resource for studying PCOs associated with breast cancer treatment by studying information posted directly by patients. We particularly focused on (i) assessing if breast cancer patients discuss health-related information on Twitter, including the self-reporting of their positive breast cancer status; (ii) developing a NLP-based social media mining pipeline for detecting self-reports via supervised classification; and (iii) analyzing health-related longitudinal information of automatically-detected users. We showed that using NLP patterns and a supervised classifier, we are able to detect breast cancer patients with high accuracy. The BERT-based classifier achieves human-like performance with an F_1_-score of 0.857 over the positive class. Qualitative analyses of the tweets retrieved from the users’ profiles revealed that they contain information relevant to PCOs, such as mental health issues, side effects of medications, and medication adherence. These findings verify the potential value of social media for studying PCOs that are rarely captured in EHRs. Our future work will focus on collecting large samples of breast cancer patients from Twitter using the methods described, and then implementing further NLP-based methods for studying breast cancer related PCOs from a large cohort.

## Data Availability

Data will be made available after peer review.

## Footnotes

¶ We use the terms ‘survivor’ and ‘patient’ interchangeably in this paper.

‖ We intend to use information from tweets labeled as F in our future studies.

